# Simple discrete-time self-exciting models can describe complex dynamic processes: a case study of COVID-19

**DOI:** 10.1101/2020.10.28.20221077

**Authors:** Raiha Browning, Deborah Sulem, Kerrie Mengersen, Vincent Rivoirard, Judith Rousseau

## Abstract

Hawkes processes are a form of self-exciting process that has been used in numerous applications, including neuroscience, seismology, and terrorism. While these self-exciting processes have a simple formulation, they are able to model incredibly complex phenomena. Traditionally Hawkes processes are a continuous-time process, however we enable these models to be applied to a wider range of problems by considering a discrete-time variant of Hawkes processes. We illustrate this through the novel coronavirus disease (COVID-19) as a substantive case study. While alternative models, such as compartmental and growth curve models, have been widely applied to the COVID-19 epidemic, the use of discrete-time Hawkes processes allows us to gain alternative insights. This paper evaluates the capability of discrete-time Hawkes processes by retrospectively modelling daily counts of deaths as two distinct phases in the progression of the COVID-19 outbreak: the initial stage of exponential growth and the subsequent decline as preventative measures become effective. We consider various countries that have been adversely affected by the epidemic, namely, Brazil, China, France, Germany, India, Italy, Spain, Sweden, the United Kingdom and the United States. These countries are all unique concerning the spread of the virus and their corresponding response measures, in particular, the types and timings of preventative actions. However, we find that this simple model is useful in accurately capturing the dynamics of the process, despite hidden interactions that are not directly modelled due to their complexity, and differences both within and between countries. The utility of this model is not confined to the current COVID-19 epidemic, rather this model could be used to explain many other complex phenomena. It is of interest to have simple models that adequately describe these complex processes with unknown dynamics. As models become more complex, a simpler representation of the process can be desirable for the sake of parsimony.

## 1 Introduction

The outbreak of the novel 2019 coronavirus disease (COVID-19) was declared a Global Health Emergency of International Concern on 30th January 2020, and pronounced a Pandemic on 11th March 2020. It has since spread rapidly with over 37 million confirmed cases and more than 1 million deaths as of 11th October 2020 [1]. Since the first reported case in December 2019, countries around the world have fought to contain the virus. In the absence of a vaccine, countries have implemented a range of non-pharmaceutical interventions and strategies to reduce the spread of the virus, from measures such as social distancing, mask-wearing and contact tracing, to complete city lockdowns and stay at home orders. These recommendations are guided by mathematical and statistical modelling to quantify the efficacy of these measures [2–9].

There is now an expansive collection of research dedicated to understanding the virus from all perspectives, including its biological, epidemiological, clinical, economic and social impacts. There is also a wealth of knowledge around prevention strategies to control the outbreak. In all of these, statistical and mathematical models are an essential aspect to gaining meaningful insights into how the virus spreads and quantifying its various impacts. A popular choice is compartmental models, with some considering the standard SIR (Susceptible-Infected-Recovered) model [10–12], and further extensions in which additional states are introduced [13–18]. As an alternative to compartmental models, others have used methods such as branching processes to capture the spread of the virus through individual networks [2, 3, 5], log-linear Poisson autoregressive models [19], and other probabilistic models of the infection cycle of the virus [20]. Various models based on growth curves have also been proposed, for example [21], [22] and [23], who use logistic, exponential and Richards growth curves respectively.

A Hawkes process [24] is a stochastic, self-exciting process in which past events influence the short-term probability of future events occurring. They are often used to explain many phenomena that exhibit self-exciting properties, including neuroscience [25–27], crime and terrorism [28–30], seismic activity [31] and social media [32]. Similarly, due to their contagious nature it is also natural to represent infectious diseases, such as the current COVID-19 pandemic, as a Hawkes process.

Hawkes processes have been successfully applied to model epidemics and infectious diseases. For example, for the Ebola outbreaks in West Africa and the Democratic Republic of Congo [33, 34], the Hawkes process is found to outperform the SEIR (Susceptible-Exposed-Infected-Recovered) mechanistic model in terms of short term prediction. Another study employs an extension of the multivariate Hawkes process to understand the transmission routes and regional connectivity for the dengue fever outbreak across regions in Australia [35]. Rocky Mountain Spotty Fever has also been modelled using a recursive Hawkes process, with the expected number of transmissions based on the current conditional intensity of the Hawkes process [36]. Moreover, [37] model invasive meningococcal disease using a spatiotemporal extension to the Hawkes process.

The spread of COVID-19 is an extremely complex process, with unknown disease dynamics and huge variations in the preventative measures and responses of different countries. We propose a parsimonious model for COVID-19 deaths, namely discrete-time Hawkes processes (DTHP) [28, 29, 38], to describe the complicated dynamics of the COVID-19 epidemic. In its original form, the Hawkes process is a continuous-time point process; however, the DTHP observes the occurrence of events at a discrete time resolution. Due to this construction, the DTHP can directly model the available data (i.e. daily counts), without artificially imputing the data onto a continuous timeline, as is generally done in studies using continuous-time Hawkes processes. We also introduce a deterministic change point in this study, since the dynamics of the spread vary abruptly as the pandemic progresses and preventative interventions are introduced.

Alternative models, such as the mechanistic and growth curve models discussed previously, primarily focus on estimating the model parameters that govern the system. Hawkes processes, however, are more detailed, as individual events and their respective occurrence times directly influence the likelihood of future events occurring. Hawkes processes also provide additional insights into the infection dynamics of diseases, through the estimation of the triggering kernel, which models the decay in infectivity through time. Hawkes processes and compartmental models are based on different mathematical principles and rely on different assumptions. However, their connection was explored by [39], who show that, via a modified variant of the Hawkes model for a particular choice of triggering kernel, the rate of events is equivalent to the infection rate in the SIR model.

### 1.1 Related Work

An approach to modelling the COVID-19 pandemic using self-exciting branching processes has been suggested by [40]. These authors employ a continuous-time Hawkes model with a nonparametric estimate of the reproduction number, *R*(*t*), the average number of secondary cases produced by a single case of the virus. Both death counts and the number of confirmed cases in the early stage of the epidemic, before April 1st, are modelled in three states of the U.S., several European countries and China. Compared to SIR and SEIR models with a fixed reproduction number, their Hawkes model with a dynamic parameter leads to lower estimates of the basic reproduction number, *R*_0_. In the same line of work, [41] consider several datasets for the state of Indiana in the early stage of the epidemic. They also compare a nonparametric estimate of the reproduction number, *R*(*t*), with an exponentially decreasing function and a step-function, and find that the estimation of *R* is very sensitive to the type of input data (i.e. deaths or cases), the data source, and the model choice. Similarly, [42] adopt a continuous-time Hawkes model with spatial covariates to model both the number of confirmed COVID-19 cases and the number of deaths, for the U.S. at the county level. This study also considers a time-varying reproduction number. Finally, [43], also use the continuous-time Hawkes process to illustrate the severity of the virus in France if no preventative action were to be taken.

Several approaches for modelling COVID-19 that incorporate change points have been proposed to capture the dynamic nature of the pandemic. [44] and [45] find that using compartmental models with time-varying infection rates, the estimated change points for Germany and South Africa, respectively, align with various government interventions in these countries. [46] do not directly estimate the change points; instead, they propose a compartmental model for Italy with piecewise model parameters partitioned into regular time intervals. Alternatively, [47] consider a combination of exponential and polynomial regression models to estimate the optimal change points for the COVID-19 outbreak in India. While these studies consider only a single country, [48] examine several countries and introduce a single stochastic change point into their compartmental model. [49] present a widespread study across 55 countries using a partially observed Markov process with piecewise transmission rates.

### 1.2 Contributions

In the current literature, the continuous-time Hawkes process requires artificial imputation of the daily count data onto a continuous time resolution, adding a significant computational burden to the implementation and adding additional, potentially unnecessary, noise to the model. We develop a multi-phase approach for the DTHP to directly model the reported daily counts of the number of deaths caused by the virus.

The dynamics of the process before and after the enactment of preventative measures and policy interventions to reduce the spread of the virus are inherently different. To the authors’ knowledge, the existing literature on the modelling of the COVID-19 pandemic using Hawkes processes consider only the early stages of the pandemic. In this work, we develop a variant of the DTHP to model the distinct phases of the COVID-19 epidemic. We modify the traditional Hawkes process to account for this change in dynamics by including a deterministic change point in the model.

Change point models for Hawkes processes have been considered in other applications [50]. However, these authors assume independence of the observed data between change points, prohibiting events that occur within a time period to influence events in future time periods. This type of model is inappropriate for this application, as the time periods are not independent. While the behaviour of the process varies between time periods, the influence of past events remains active in the memory of the process. Thus, the baseline parameters become artificially inflated if events from different time periods are assumed to be independent.

In particular for the COVID-19 epidemic, while other studies directly estimate the change points or partition the timeline into regular intervals to reflect the evolving dynamics of the epidemic, we propose a simple method that incorporates only a single, fixed change point. We do not attempt to estimate the change point for our model, as different interventions have been introduced in each country, with varying levels of restrictions. Furthermore, the delays before tangible results are observed, in addition to the complex and hidden interactions underlying the process, complicate the interpretation of estimated change points. We instead opt for a consistent and simplistic definition of the change point for each country.

We illustrate in this study how a simple model can be used to describe exceedingly complex natural phenomena such as epidemics, and in particular the COVID-19 pandemic. Although it is the same underlying phenomenon, all countries are unique concerning the spread of the virus and the resultant response measures. Our simple model can capture these dynamics. Additionally, while many other studies consider small-scale regions, such as individual counties in the U.S., we are also able to gain insights into the dynamics of the process at a higher-level across entire countries.

### 1.3 Outline

In Section 2.1, a general form of the DTHP is defined, and contrasted with its continuous-time equivalent. Section 2.2 follows by introducing the particular model used in this analysis for modelling COVID-19, incorporating a change point into the construction of the DTHP. We provide a brief description of the data and inference methods in Sections 2.3 and 2.4 respectively. Section 3 then presents the results for the ten countries of interest, followed by a discussion in Section 4 and final conclusions in Section 5.

## 2 Methods

### 2.1 Discrete-time Hawkes process

The discrete-time Hawkes process is a self-exciting stochastic process whereby events occur at regular intervals on a discrete-time scale. It follows a similar construction to the continuous-time Hawkes process [24]. The conditional intensity function *λ* (*t*) characterises a Hawkes process, and herein lies the difference between the continuous-time and discrete-time variants. For the DTHP, *λ* (*t*) represents the expected number of events that occur at time interval *t*, conditionally on the past. In contrast, for the continuous-time Hawkes process, *λ* (*t*) is the instantaneous rate of an event occurring at time *t*. The DTHP model also has an extra layer of flexibility compared to its continuous-time counterpart as the underlying data generating process can be selected as any counting distribution with conditional mean *λ* (*t*).

Consider a linear univariate discrete-time Hawkes process *N*, where *N* (*t*) represents the number of events up to time interval *t. N* (*t*) is dependent on the history of events up to but not including time *t*, denoted by *H*_*t*−1_ = *{y*_*s*_ : *s* ≤ *t* − 1*}*, where *y*_*s*_ represents the observed number of events in a given time interval *s*. Furthermore, *N* (*t*) − *N* (*t* − 1) represents the number of event occurrences at time *t*, and thus,

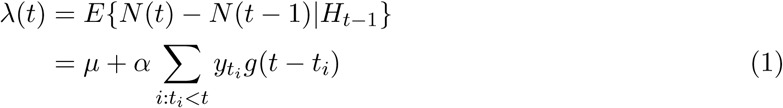

where *μ* represents the baseline mean of the process and the second term represents the self-exciting component of the Hawkes process, describing the expected number of events during a particular interval *t* given previous events. The triggering kernel *g*(*t*− *t*_*i*_) describes the influence of past events on the intensity of the process, given the time elapsed since event *i*, where *t* > *t*_*i*_. In this study, we specify the triggering kernel to be a proper probability mass function with strictly positive support. Thus one can interpret the non-negative magnitude parameter *α* ∈ ℝ _≥0_ as the expected number of subsequent events produced by a single event [29].

### 2.2 Model

Daily counts of the reported number of deaths of the novel coronavirus COVID-19 are modelled using the discrete-time Hawkes process, where the number of events observed on day *t*, namely *y*_*t*_, are distributed according to the random variable, *Y* (*t*), which has conditional mean *E*(*Y* (*t*)|*H*_*t*−1_) = *λ* >(*t*) as defined in (1). In this analysis *Y* (*t*) is assumed Poisson distributed, thus *Y* (*t*) ∼ *𝒫* (*λ* (*t*)). The Poisson distribution is selected as it has an intuitive interpretation regarding the generation of daily death counts on a given day and also to align with the assumptions of the continuous-time Hawkes process, whereby the underlying generative model follows a Poisson process. Thus, for the proposed DTHP model, the probability that day *t* has *y* events is,

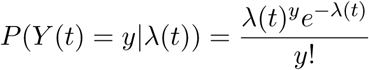

The conditional intensity function *λ* (*t*) is altered from (1) to allow for change points in the process, since the DTHP with fixed parameters is unable to capture the complex dynamics for an epidemic of this scale. The parameters of the DTHP implicitly incorporate environmental and social characteristics that are significant for the spread of the disease, and these characteristics change after preventative measures are introduced. Thus, if the dynamic nature of the epidemic is not taken into account, the model averages the estimated parameters, combining the effects of the initial explosive phase of the pandemic with the downward trend that follows after the implementation of preventative measures.

Thus, we retrospectively define a single change point at time *T*_1_, where *T*_1_ is the maximum value of deaths, to capture the different dynamics of the epidemic at two distinct stages of the outbreak. Intuitively, the time series before *T*_1_ represents the first stage in the epidemic where the virus is spreading rapidly, and the time series after *T*_1_ represents the process following the introduction of preventative measures and policies, and when cases have started to decline.

The conditional intensity function before *T*_1_ is calculated using one set of model parameters, (*μ*_1_, *α*_1, 1_). After *T*_1_, the intensity function is calculated using a new set of parameters, (*μ*_2_, *α*_2_, *β*_2_) for the second phase in the epidemic. Thus for one change point at time *T*_1_, *λ* (*t*) is given by,

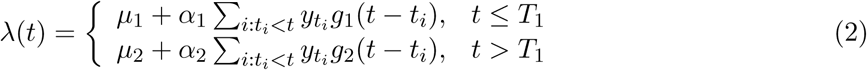

It is straightforward to extend Equation (2) to allow for additional change points. However, this did not appear to be justified for our analysis and indeed might limit model performance due to lack of information in shorter time series.

The triggering kernel *g*(*t* − *t*_*i*_) is selected as a geometric excitation kernel, *g*(*i*; *β*) = *β* (1 − *β*)^*i*−1^. We choose the geometric kernel to resemble the exponential distribution, which is one of the most commonly used triggering kernels for continuous-time Hawkes processes.

For a time series of *T* days and a given country, the log-likelihood function for this DTHP model with retrospective change point, *T*_1_, up to an additive constant *K*, is then,

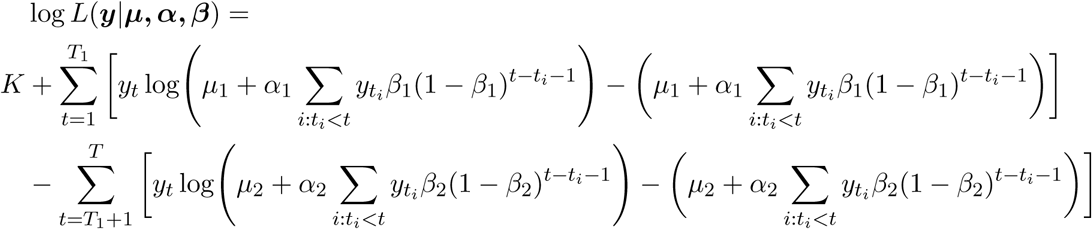

### 2.3 Data

We use data gathered by the Johns Hopkins University [51] in this work. These data come in the form of daily counts of confirmed cases or deaths by country and region. In this analysis, the number of daily reported deaths for a selection of countries, namely Brazil, China, France, Germany, India, Italy, Spain, Sweden, the United Kingdom and the United States, are considered. We select these countries to represent a global sample of countries that have been adversely affected by the coronavirus outbreak. It is important to note that the definition of deaths due to COVID-19 varies between countries. These differences are ignored in our modelling.

The reported number of deaths was considered a more reliable response variable than the reported number of cases. This is due to data issues that can arise when considering the number of confirmed cases, such as lack of testing or differing testing rates between countries, differences in definitions and differences in the timing for reporting of cases. Additionally, to mitigate the effect of systematic influences in reporting, such as lower reporting on weekends [44], the data is smoothed over a rolling window of seven days. The start of the observation window, *t*_1_, for each country is defined as the time the number of deaths exceeds ten. Fig 1 shows the smoothed volume of daily deaths for the countries under consideration up to 25th July 2020.

**Fig 1.**
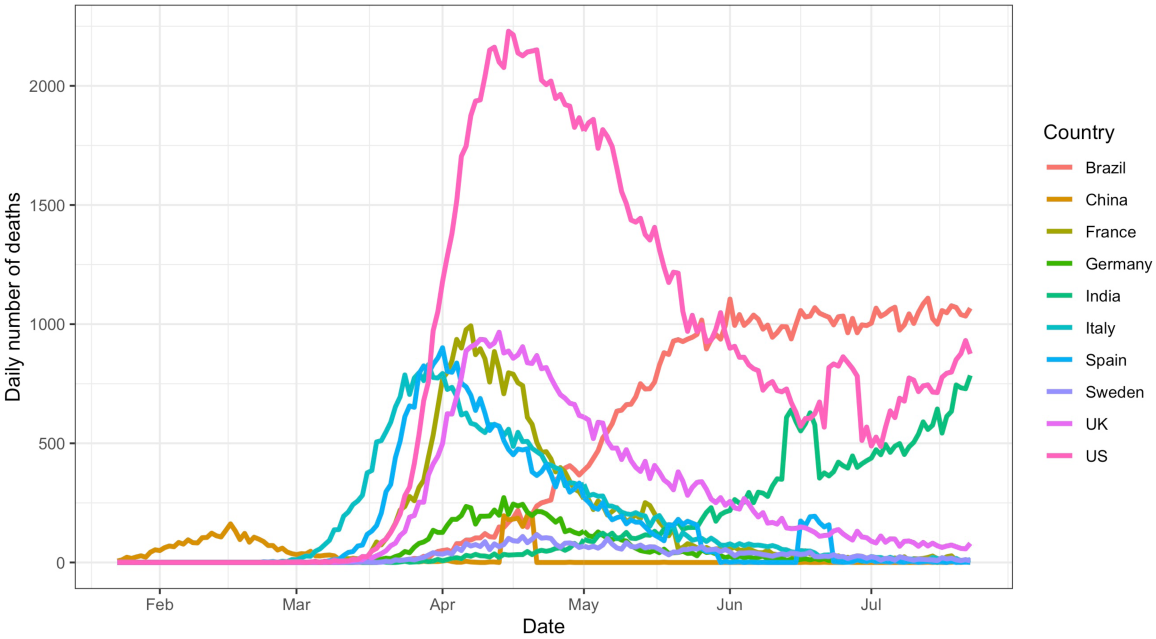
Observed data. Daily volume of deaths due to COVID-19 for the countries selected in this analysis.

We define the change point, *T*_1_, as the time where the maximum number of deaths occurs, for the countries with sufficient data in the downward phase of the epidemic by 25th July 2020. Where there is insufficient evidence for the downward trend, for example, in India and Brazil, no change point is introduced, and only a single phase is modelled. Moreover, the trend for Brazil shows evidence of the curve flattening; however, there is insufficient data for this second phase. Thus the end of the observation window for Brazil is fixed on 1st June 2020. Additionally, as China, India, Spain and the United States experienced large deviations from the current trend towards the end of the observed data, earlier endpoints of 13th April 2020, 12th June 2020, 15th June 2020 and 21st June 2020 were imposed respectively. This avoids the anomalous spikes at the end of these series, since it was not clear whether these aberrations were real or due to reporting definitions or other errors. The endpoint for the remaining countries is set as 25th July 2020.

### 2.4 Parameter Inference

Parameter estimation is undertaken using Bayesian methods. We consider a range of prior choices for the baseline parameters *μ*_1_ and *μ*_2_, and perform leave-future-out cross validation with Pareto smoothed importance sampling [52] to assess the performance of each prior choice. The priors considered are,

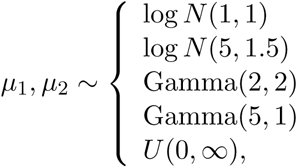

where the first term of the log-normal priors represents the mean of the random variable itself, as opposed to the mean of the variable’s natural logarithm.

Cross validation with Pareto smoothed importance sampling relies on the expected log predictive density (ELPD), for which a larger value indicates a better model fit. We calculate the ELPD in each country for each of the baseline parameter prior choices, and these results are provided in the supplementary material. Based on this analysis, there is no obvious choice of prior that consistently outperforms the rest for each country. On the contrary, the difference in the ELPD is marginal between priors. The remainder of this paper presents the results for *μ*_1_, *μ*_2_ *∼* Gamma(5, 1), as this is most frequently the highest ELPD, and if not the maximum, is generally very comparable.

Flat priors are selected for *α*_1_, *α*_2_, *β*_1_ and *β*_2_ such that,

- 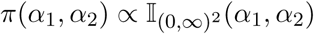
- *β*_1_, *β*_2_ *∼ U* (0, 1)

A Metropolis-adjusted Langevin step [53] is used to jointly update *α*_1_ and *β*_1_, and also to jointly update *α*_2_ and *β*_2_. Denoting the parameters at iteration *t* by *α* ^(*t*)^, *β* ^(*t*)^, the proposals *α*^*^, *β*^*^ are simulated from,

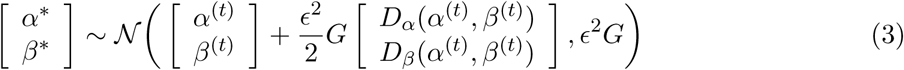

where *D*_*α*_ (.) and *D*_*β*_ (.) are the gradients of log *L* with respect to *β* and respectively, *ϵ* is a pre-conditioning matrix accounting for covariance between parameters and *ϵ* is the step size in the Metropolis-adjusted Langevin algorithm.

The MCMC chain was run for 60,000 iterations discarding the first 20,000. The pre-conditioning matrix *G* was taken as the covariance matrix from an implementation of the standard Metropolis-Hastings algorithm for each country. The R code and data required to replicate this study are available on Github (https://github.com/RaihaTuiTaura/covid-hawkes-paper).

## 3 Results

Fig 2 presents the 95% posterior intervals around the estimated conditional intensity function *λ* (*t*) against the observed data for each country. The estimated intensity function on day *t*, represents the expected number of events on day *t* and very closely follows the observed number of deaths. It is also extremely reactive to minor deviations from the observed trend, and more volatile times in the observed data result in wider posterior intervals to account for increased uncertainty in the trend of the data.

**Fig 2.**
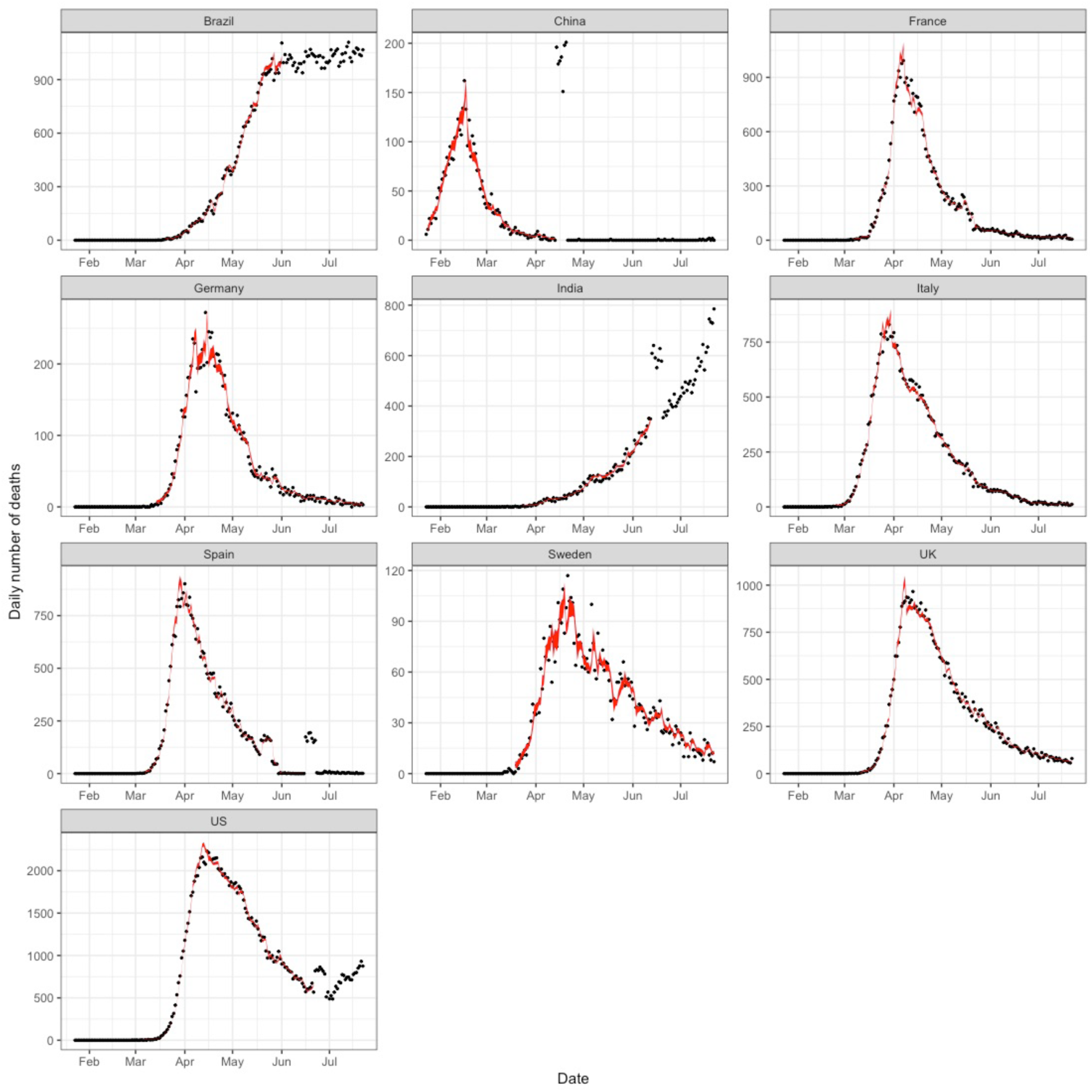
Observed deaths versus estimated deaths. The observed number of deaths (black dots) compared to the 95% posterior interval for the estimated expected number of events, i.e. *λ* (*t*) (solid red ribbon).

Diagnostic plots, including MCMC trace plots, autocorrelation between the MCMC samples and pairwise correlation between parameters were examined and suggest the algorithm has converged. Further details on the posterior distributions of the model parameters, convergence and model diagnostics are provided in the supplementary material.

Tables 1-3 present the posterior median and corresponding 80% posterior intervals for the model parameters. Further details for the other baseline parameter priors considered can be found in the supplementary material. In most countries, the posterior interval for *μ*_2_ is consistently lower than *μ*_1_, indicating a reduction in the baseline rate of events from the beginning to later stages of the epidemic. The exception to this is the U.S. The results for the U.S. are highly sensitive to the prior choice; thus, wider priors return higher posterior estimates than expected when compared to other countries. In an earlier analysis, this behaviour was also prevalent for Sweden and the U.K., although it disappeared when considering a longer time series. This implies that there may be insufficient information in the data for the U.S. to reliably learn the model parameters for the second phase. However, without alternative data, it is not possible to improve modelling for the U.S. by considering a longer time series. This is due to a large anomaly at the end of the series, as discussed in Section 2.3. Nonetheless, it highlights the importance of having sufficient training data and being cautious when interpreting parameter estimates.

**Table 1.** Phase 1 versus Phase 2 median and 80% intervals for baseline parameters, *μ*_1_ and *μ*_2_

| Country | $\mu_1$ | $\mu_2$ |
| --- | --- | --- |
| Italy | 4.39 (3.18,5.71) | 1.17 (0.69,1.8) |
| France | 4.57 (3.38,5.91) | 1.57 (0.97,2.28) |
| Spain | 5.78 (4.06,7.6) | 0.49 (0.28,0.76) |
| Germany | 4.17 (2.89,5.54) | 0.95 (0.59,1.39) |
| Sweden | 4.05 (2.88,5.44) | 1.79 (1.05,2.68) |
| U.K. | 4.51 (3.08,6) | 2.42 (1.32,3.75) |
| U.S. | 4.08 (3.13,5.15) | 4.1 (2.16,7.12) |
| China | 8.92 (6.29,11.73) | 0.82 (0.48,1.22) |
| Brazil | 4.18 (2.98,5.52) | - |
| India | 2.81 (2.02,3.72) | - |

**Table 2.** Phase 1 versus Phase 2 median and 80% intervals for magnitude parameters, *α*_1_ and *α*_2_

| Country | $\alpha_1$ | $\alpha_2$ |
| --- | --- | --- |
| Italy | 1.07 (1.05,1.09) | 0.94 (0.93,0.95) |
| France | 1.1 (1.08,1.11) | 0.92 (0.91,0.93) |
| Spain | 1.11 (1.09,1.13) | 0.96 (0.95,0.97) |
| Germany | 1.06 (1.03,1.09) | 0.91 (0.89,0.93) |
| Sweden | 1.07 (1.01,1.13) | 0.92 (0.89,0.95) |
| UK | 1.14 (1.11,1.17) | 0.95 (0.95,0.96) |
| US | 1.07 (1.06,1.07) | 0.97 (0.97,0.98) |
| China | 1.07 (1.01,1.15) | 0.8 (0.76,0.84) |
| Brazil | 1.03 (1.02,1.04) | - |
| India | 1.1 (1.07,1.13) | - |

**Table 3.** Phase 1 versus Phase 2 median and 80% intervals for triggering kernel parameters, *β*_1_ and *β*_2_ and the means of their respective geometric distributions, 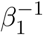 and 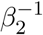

| Country | $\beta_1$ | $\beta_2$ | $\beta_1^{-1}$ | $\beta_2^{-1}$ |
| --- | --- | --- | --- | --- |
| Italy | 0.88 (0.8,0.95) | 0.55 (0.48,0.63) | 1.136 (1.053,1.25) | 1.818 (1.587,2.083) |
| France | 0.97 (0.92,0.99) | 0.64 (0.58,0.7) | 1.031 (1.01,1.087) | 1.562 (1.429,1.724) |
| Spain | 0.96 (0.9,0.99) | 0.91 (0.85,0.95) | 1.042 (1.01,1.111) | 1.099 (1.053,1.176) |
| Germany | 0.65 (0.57,0.75) | 0.51 (0.45,0.59) | 1.538 (1.333,1.754) | 1.961 (1.695,2.222) |
| Sweden | 0.42 (0.32,0.54) | 0.5 (0.39,0.62) | 2.381 (1.852,3.125) | 2 (1.613,2.564) |
| UK | 0.79 (0.68,0.91) | 0.56 (0.5,0.62) | 1.266 (1.099,1.471) | 1.786 (1.613,2) |
| US | 0.99 (0.98,1) | 0.77 (0.66,0.89) | 1.01 (1,1.02) | 1.299 (1.124,1.515) |
| China | 0.4 (0.28,0.56) | 0.43 (0.35,0.54) | 2.5 (1.786,3.571) | 2.326 (1.852,2.857) |
| Brazil | 0.83 (0.73,0.93) | - | 1.205 (1.075,1.37) | - |
| India | 0.33 (0.26,0.41) | - | 3.03 (2.439,3.846) | - |

The magnitude parameter in the second phase, *α*_2_, is also consistently lower than the parameter for the first phase, *α*_1_. With a posterior probability (greater than 80%), it can be said for all countries that *α*_1_ > 1 and *α*_2_ *<* 1. This implies the process is explosive before the change point and becomes stationary after the change point, likely driven by the introduction of interventions to reduce the rate of infection.

The parameters for the geometric triggering kernel, *β*_1_ and *β*_2_, are similar for Sweden and China. However, for the remaining countries where two phases are considered, the kernel parameter for the first phase, *β*_1_, is larger than *β*_2_, indicating that the self-excitation has a longer memory in the second phase. For reference, *β* = 0.4 in the geometric kernel corresponds to an average of 2.5 days for the self-excitation, with the majority of the mass occurring within one week, whereas *β* = 0.9 is shorter, corresponding to an average self-excitation of just over 1 day with approximately 2 days of total memory.

### 3.1 Model Fit

Several measures are used to assess model fit. First, the model’s capability to interpolate missing data is evaluated. Then in-sample and out-of-sample posterior predictive checks are considered. The purpose of prediction in this study is to assess model fit and to discover what can be learned about the process retrospectively.

The first measure of model fit considers how accurately the model can recover missing data. We randomly remove 10% of observations across the entire time series and treat the missing data as parameters in the model to estimate. Table 4 describes the number of interpolated data points for which the observed value lies within both the 95% and 80% credible intervals (CrI) of the posterior distributions for the missing data. The proportion of data points correctly interpolated is generally high when considering the 95% credible intervals. This reduces when considering the 80% interval, however, is still high for most countries, capturing at least half of the missing data points. The exception to this is the U.S., with just less than half of the missing data points accurately interpolated.

**Table 4.** Number of missing data points with actual value within 95% and 80% CrIs, out of the total number of missing data points.

| Country | 95% CrI (average) | 80% CrI (average) |
| --- | --- | --- |
| France | 11/14 | 7.4/14 |
| Italy | 13/15 | 11/15 |
| Germany | 13.4/14 | 10.2/14 |
| Spain | 8/11 | 6.2/11 |
| Sweden | 12.6/13 | 10.4/13 |
| U.K. | 11.8/14 | 9.2/14 |
| China | 8.6/9 | 7.2/9 |
| U.S. | 8.6/11 | 5.4/11 |
| Brazil | 6.6/8 | 4.6/8 |
| India | 7.8/9 | 6.8/9 |

Prediction is a difficult task, particularly for complex phenomena such as the COVID-19 pandemic. For this particular model, more recent events have a larger impact on the intensity of the process, and thus the model is very reactive to recent behaviours. Thus prediction performed at a time where abnormal behaviour is occurring will be highly uncertain and often unreliable. Moreover, a prediction is only realistic in the short term and generally only at times where there is no evidence of abnormal behaviour. This is consistent with other models in the literature [33,34,54–56]. Thus we consider in-sample and out-of-sample posterior predictive checks in this study as a measure of model fit only.

In-sample prediction is performed by generating sample paths of the process for the range of model parameters obtained and comparing these to the observed time series. In particular, a random selection of posterior samples is taken, and the entire time series is simulated from these draws. The posterior predictive intervals from these simulations compared to the observed data are given in Fig 3. In general, the intervals for these simulations encapsulate or are very close to the observed data, however, they can be extremely wide and often underestimate the volume of events in the initial phase of the outbreak. This is likely due to variation in the assumed Poisson data generating distribution, and relatively wide priors on the baseline parameters for the first phase, resulting in a wide range of possible sample paths. Additionally, these sample paths did not adequately capture the observed trend in the U.S. However, we find that including the data from the first phase in the model and predicting the second phase results in improved accuracy of the posterior predictive intervals for all countries. These results are presented in Fig 4.

**Fig 3.**
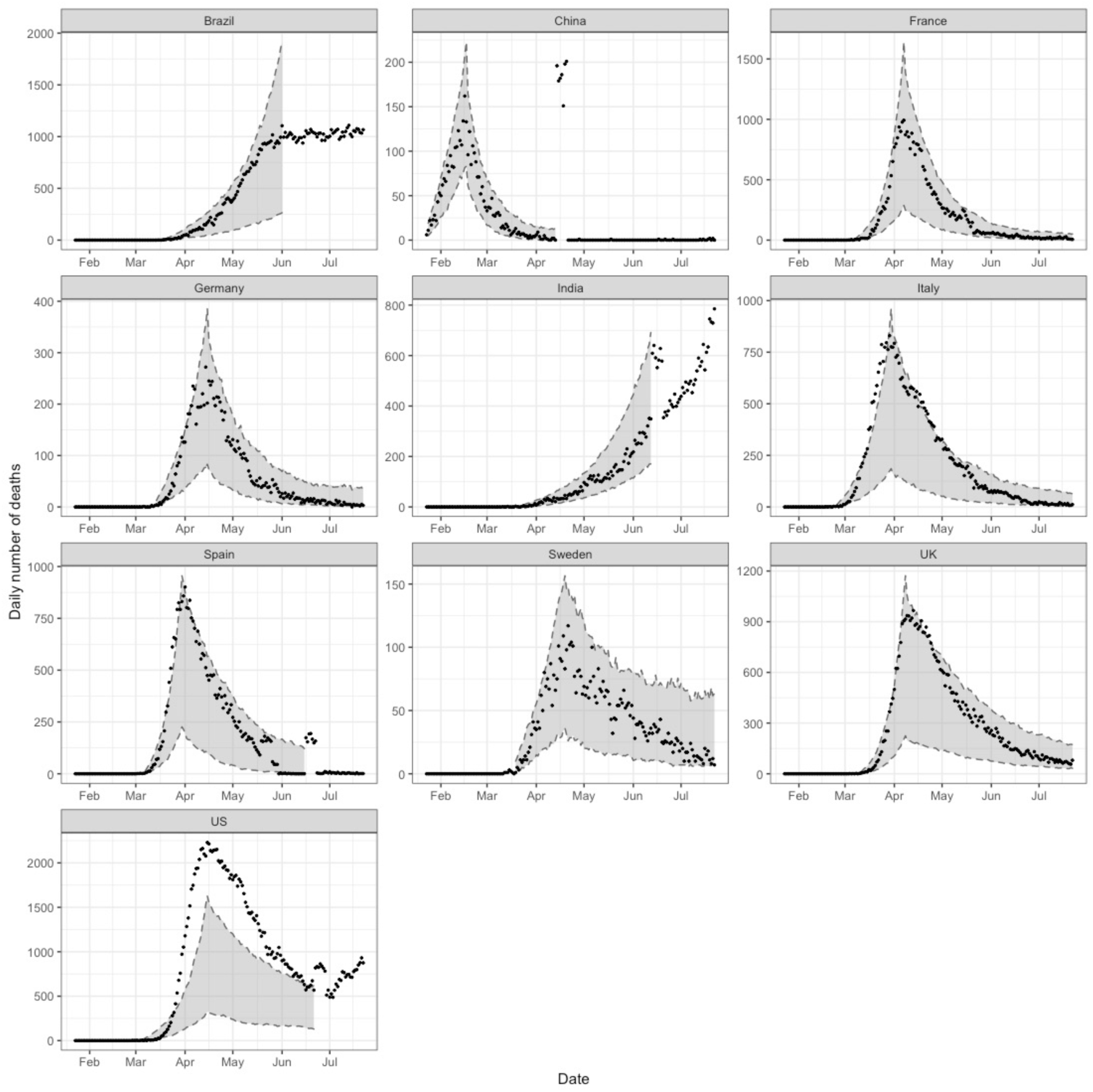
In-sample validation. The observed number of deaths (black dots) compared to the 95% posterior predictive interval for the estimated expected number of events, i.e. *λ* (*t*) (grey ribbon).

**Fig 4.**
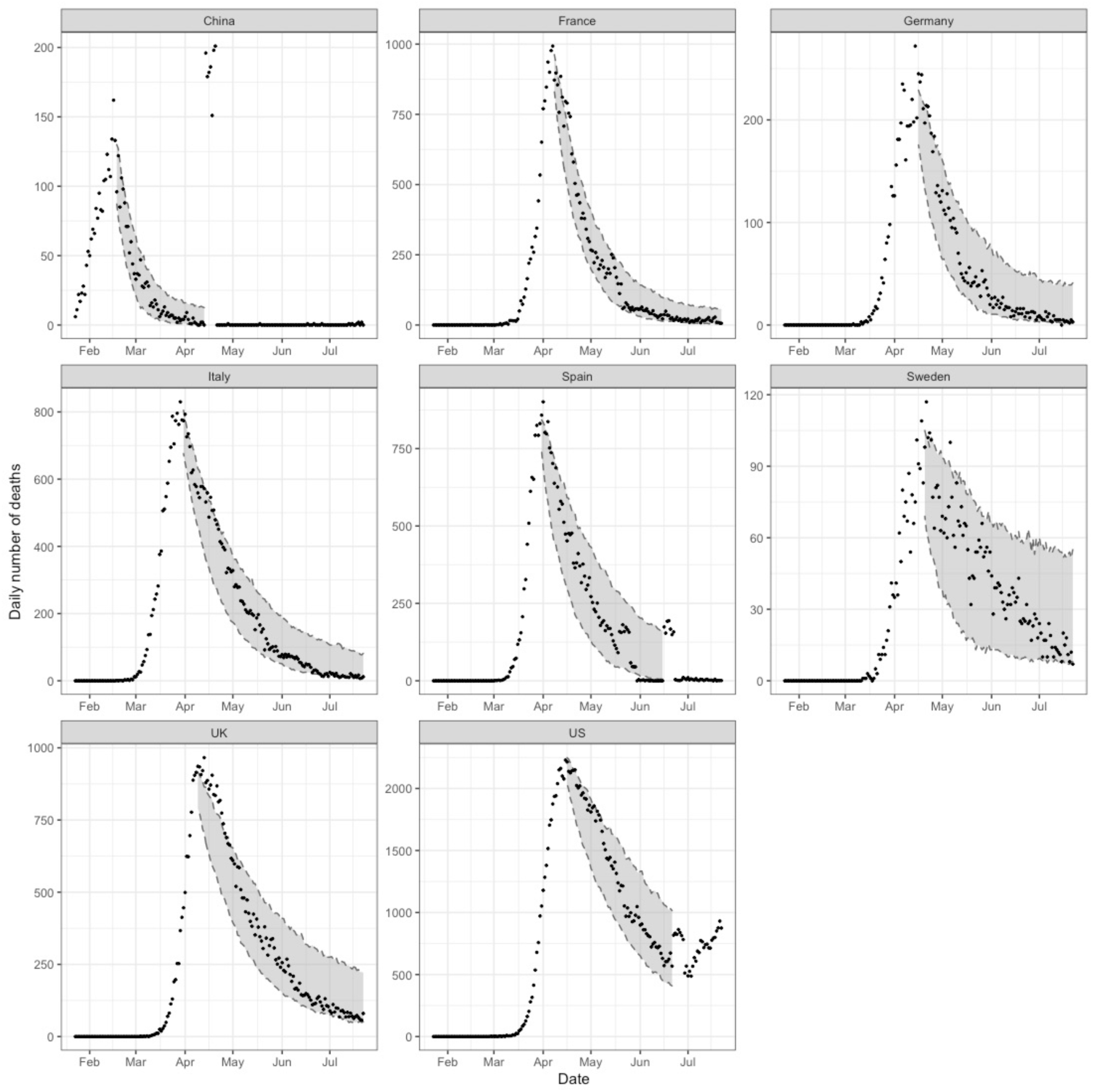
In-sample validation, conditioned on data from the first phase. The observed number of deaths (black dots) compared to the 95% posterior predictive interval for the estimated expected number of events, i.e. *λ* (*t*) (grey ribbon).

Out-of-sample (O.O.S.) validation is also performed for each country as a measure of model fit. First, we consider the initial phase of the epidemic before the change point. The model is trained on data from the first 15 days of the sample, followed by a 5-day O.O.S. prediction. We then repeat this process, increasing the length of the training period by 5 days until the change point. As shown in Fig 5, these predictions are reliable only in the short term, and become more unreliable as the end of the first phase approaches. The first phase predictions grow exponentially and quickly surpass the actual growth of the process, as the observed curve flattens due to the effects of preventative measures that have been implemented.

**Fig 5.**
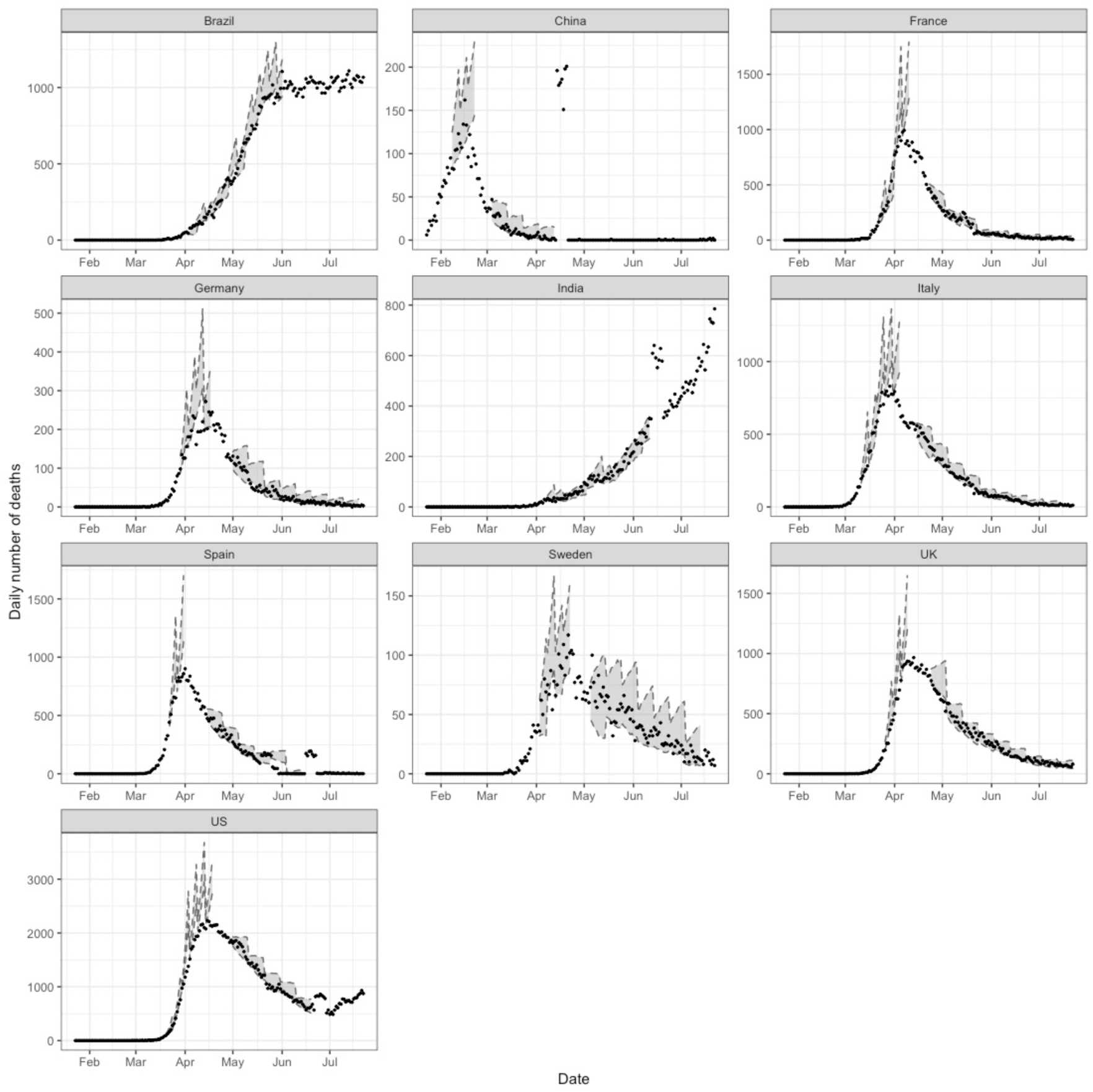
Out-of-sample validation. The observed number of deaths (black dots) compared to the 95% posterior predictive interval for the estimated expected number of events, i.e. *λ* (*t*) using various training datasets (grey ribbons).

O.O.S. prediction is also considered for the second phase of the model, after the change point. We first train the model on data from the first phase and 15 days of the second phase. We then repeat the same procedure as described above with 10-day O.O.S. predictions. The downward trajectory of the infection cycle is more stable than the upward trajectory, so we consider a longer prediction duration. The posterior predictive intervals are generally very accurate for all countries, as seen in Fig 5. Compared to the O.O.S. validation performed for the first phase, the improvements in accuracy observed in the second phase are likely due to the stationarity of the process in the second phase, resulting in more predictable trends. For both phases, the accuracy of O.O.S. predictions depends on the endpoint of the training period for the model, and the type of behaviour preceding any predictions.

While we do not attempt to predict the course of the epidemic in this study, we do find that O.O.S. predictions may indicate when the peak in the number of events is approaching. This could be useful in countries that have not yet experienced a decline in the number of daily events, for example, Brazil and India in this study. Posterior predictive intervals that surpass the growth rate in the observed data indicate, and could pre-empt, the downward phase of the epidemic. Conversely, where the predictive intervals do encapsulate the observed data, it is unlikely that the peak is being approached. This is evident in Fig 5, where the curve for Brazil is flattening, resulting in unreliable O.O.S. predictions, compared to the more reliable predictions in India due to the strong upward trend.

## 4 Discussion

Infectious diseases have previously been studied using Hawkes processes. However, the scale, severity and uncertainty of the current COVID-19 pandemic make it a very challenging problem, providing a unique opportunity to evaluate the capacity of Hawkes processes in describing an incredibly complex process. Another source of complexity arises from the definition of what constitutes a COVID-19 death, which differs between countries. This analysis finds that by modifying the DTHP to incorporate a change point, our model can adequately capture the overall process as two distinct phases, while quickly reacting to and accommodating for some level of abnormal behaviour.

The findings of this work can also quantify the dynamics of these two distinct phases in the pandemic. Our results show that for the baseline parameters, the background rate in the second phase, *μ*_2_, is lower than that for the first phase, *μ*_1_. This is analogous to a reduction in the baseline level of exogenous events, possibly related to reduced travel and general mobility. Another factor could be increased levels of community transmission, affecting the self-exciting component of the intensity function, and thus placing less emphasis on the baseline component. The exception to this is the U.S., for the reasons stated in previous sections. The baseline parameter could also be affected by the definition of a reported COVID-19 death, as this differs between countries. For example, when the criteria for reporting a death excludes cases where the person suffers from other illnesses in addition to the virus, this could result in an inflated baseline rate, as secondary events from unreported cases could be present in the data.

Our results for the magnitude parameters show, with a high degree of certainty, that for the first phase *α*_1_ is greater than 1, and for the second phase *α*_2_ is less than 1. This exhibits the distinct differences between phases, as a magnitude parameter greater than 1 indicates the process itself is non-stationary, and similarly a magnitude parameter less than 1 suggests a stationary process. We discuss below the similarities between the magnitude parameters in our model and the reproduction number in standard epidemiological models.

The triggering kernel parameter in the first phase, *β*_1_, is higher than that for the second phase, namely *β*_2_, for all countries except Sweden and China. This could suggest that in later stages of the epidemic when preventative measures have been implemented, the time between transmission is longer, as there is less opportunity for transmission. The two exceptions to this, Sweden and China, are on opposite ends of this spectrum. While China enforced very strict lockdown and quarantine requirements, Sweden adopted a soft approach to lockdown.

Throughout this analysis, we have found difficulty in fitting the proposed model for the U.S. In particular, the posterior estimates for the baseline parameter are uncertain as they are heavily influenced by the prior choice. Additionally, in-sample posterior predictive checks found that the sample paths produced by the estimated model parameters do not resemble the observed trend. We consider the U.S. an anomaly, as their response to the virus by the relevant state-level authorities varied widely between states. While this is also true to an extent for other countries, the heterogeneity across the country was arguably more significant for the U.S., implying that the proposed model may need to be applied at a more granular level of regions to obtain more reliable results. Despite this model being able to accurately capture the dynamics of this complex process, some limitations and extensions could be considered. As the epidemic is still ongoing, new data is becoming available each day, and the model must be re-fit and re-tuned each time the data is updated. Additional change points can also be considered when there are significant changes in the trend, such as second waves of infection. An algorithm with automatic selection of the number of change points and their respective locations could also be considered. However, additional change points need to be determined carefully as the length of time between each change point must be sufficient. Another consideration is a Bayesian nonparametric spline [57], providing time-varying parameters through flexible splines. However, the identifiability and existence of this model would need to be established. One could also consider different triggering kernels, including nonparametric kernels in order to improve the flexibility of the model.

Our model considers only the infected population, as opposed to standard epidemiological models that differentiate the population into several groups depending on their infection status, for example, the SIR model. It is helpful to consider a stochastic variation of the SIR model as a bivariate Poisson process, comprised of infection and recovery events, to compare the two frameworks. Infection events are then governed by a Poisson process where the rate is based on the transmission rate and the current size of the susceptible and infected populations, corresponding to the rate of infection in the deterministic SIR model. Our model differs as we consider a discrete time scale, the daily number of events is Poisson-distributed and, conditioned on past events, the rate of events each day is given by (2).

The reproduction number, defined as the number of secondary infections from a single case, is a crucial parameter in epidemiological models. Similarly, the magnitude parameters in our model, *α*_1_ and *α*_2_, also represent the expected number of secondary cases caused by a single parent event. While their respective interpretations are similar at a superficial level, *α*_1_ and *α*_2_ are not directly comparable to reproduction numbers in epidemiological models, due to differences in model assumptions and the underlying mathematical frameworks. Furthermore, the magnitude parameters in our model do not provide the same information as reproduction numbers. Examples include the level of herd immunity that will bring the virus under control, and the proportion of new infections that must be prevented to change the trend of events from increasing to decreasing [58]. Other key epidemiological parameters are generation times and serial intervals, which describe the time between infection and development of symptoms, respectively, for a pair of individuals. Our model does not capture this type of information, as we do not consider the relationship between specific pairs of individuals. As a result, it is not possible to obtain parameters such as growth rates, which are often of interest in epidemiological models. However, we can gain insight into an alternative temporal aspect of the contagion. The geometric triggering kernel in our model describes how the probability of contagion changes as time elapses. More precisely, we can determine, for a given day, the influence of past events on the expected number of events for that day.

## 5 Conclusions

The utility of our model is not restricted to the current coronavirus epidemic, and could be used as a simple model to describe a much broader range of complex phenomena. We have demonstrated through this study that the proposed model is a simple, yet powerful tool for explaining an incredibly complex process. In general, models that attempt to describe complex processes can become increasingly complicated, as more intricate details are embedded and accounted for in the modelling. Thus having a parsimonious model that is flexible enough to competently capture the dynamics of a complex process, without adding too much additional complexity, is very desirable.

In particular for the current pandemic, this study shows that our simple discrete-time Hawkes process can capture the dynamics for different countries, despite the complexities involved with each country’s unique response to the virus. The same underlying biological process is affecting countries in different ways, and there is a significant difference in the impact and severity of the pandemic across different countries. Additionally, the actions that have been taken to stop the spread, and the timing of these also vary widely. These different behaviours between countries mean that the evolution of the pandemic for an individual country is very intricate within itself, and involves many unseen and complex hidden interactions that we cannot model directly. However, the proposed model, while being very simple, can capture these trends surprisingly well.

To adequately model the entire course of the pandemic, we find that we must make provisions as there are multiple distinct phases. Initially, there is exponential growth as the virus spreads, followed by a period of reduced infection rates as actions are taken to slow the spread. These distinct behavioural differences throughout the evolution of the epidemic must be acknowledged, as a single DTHP applied to the entire time series provides uninformative and uninterpretable parameter estimates. Hence a model that accounts for these different phases, such as the model presented in this work, is required.

Fitting a DTHP to the epidemic has led to some other unique insights. Our results show that a discrete-time model is appropriate for this application, avoiding unnecessary computational burden as well as additional noise due to artificial data imputation, as is required for the continuous-time model. This model also provides to an extent, interpretable parameters and an indication of the changing dynamics between distinct phases of the pandemic. We show that despite unique circumstances for individual countries, including the type and timing of non-pharmaceutical interventions, population demographics, and the overall impact of the virus, the model is flexible and can also accomodate some level of volatility in the data. Furthermore, one of the most surprising outcomes of this analysis is that, at the country level, a very simple DTHP model fits remarkably well to the number of deaths, thus capturing the dynamics of the COVID-19 pandemic.

## Data Availability

Data is available in the public domain.

https://github.com/CSSEGISandData/COVID-19

## Acknowledgements

The authors are grateful to Dr Gentry White, for helpful advice on modelling discrete-time Hawkes processes in the early stages of this project.

